## Supplemental Table 1 for "Exploring Pathways to Compulsory Detention and Ways to Prevent Repeat Compulsory Detentions; Clinician Perspectives"

| **Theme** | **Service User Level Factors Influencing Risk of Being Compulsorily Detained** | | |
| --- | --- | --- | --- |
| **Sub Themes:** | **High Levels Of Risk** | **Previous/Underlying Trauma** | **Service User Perceptions of Their Own Mental Wellbeing and/or Diagnosis** |
| **Additional Quotes** | “So, it was about risk. It was about his safety. Not his risk to others, his risk to himself, because he wasn’t able to see how inadequate his solution was.” [P15]  “Because in this role you’re always weighing up the possible long-term detrimental effects of repeated admissions with a person’s short term wellbeing and safety. And that’s a constant, kind of, yes, dilemma, I suppose, clinically and ethically. That’s why the work in a way is, kind of, interesting and challenging, but also, yes, anxiety provoking and stressful as well at times because, you know, it’s a person’s life, care, and wellbeing that’s all at stake.” [P4]  “Sometimes we find it difficult to separate a historical risk from a current risk and that results in people being detained more often….” [P17]  “High level of acute risk in terms of threat to her life, very chronic presentation, a sense of hopelessness and desperation all in how she was presenting.” [P1]  “He’d given away all his belongings. He had been keen to move away from supported accommodation to find a flat with friends. We know he doesn’t have friends, so worried about his vulnerability.” [P15] | “.. their collective cultural, the historical trauma that they may carry, and what involving the police might mean to a certain group of people, compared to another group of people, and what being detained, being locked up… I think some of these groups may be more likely to be referred to PICU straightaway, to be on higher doses of anti-psychotics.” [P20]  “She appeared to be in a kind of, a crisis in that sense, an emotional crisis, and that was linked to some anniversary dates around traumatic events such as the loss of a child, so a very significant loss to that person as well as, you know, as well as other stresses around other children who have been removed from her care.” [P4]  “This is somebody who has had a really difficult experience growing up, and experience of the care system.” [P8] | “They didn’t have the capacity to weigh up the … or engage in the assessment really and weigh up the treatment options.” [P14]  “…his understanding of his situation wasn’t full. He was seeing things from a particular perspective, but he wasn’t really taking account of the trouble that he was in.” [P16]  “This morning, I had a meeting with social services about one of my pregnant patients. There are no insights, she doesn’t have insights. If you ask her, she will tell you, “I don’t have a mental illness, I’m just [speaking to Allah].” She’s on clozapine, which is an antipsychotic. She has had an admission of six months.” [P15]  “People’s understanding of their illness, their insight into their illness and their insight into what will happen is really, really key as to whether you end up going down the Mental Health Act route.” [P17] |

| **Theme** | **Service User Level Factors Influencing Risk of Being Compulsorily Detained** | | | |
| --- | --- | --- | --- | --- |
| **Sub Themes:** | **Non-Adherence to Medication** | **Lack Of Support from The Service User’s Family and Networks** | **Disadvantage and Discrimination** | **Cultural Attitudes to Mental Illness And Services** |
| **Additional Quotes** | “We do see frequently that people relapse because they stop taking their medication. And that may be because they’ve got better, they feel better, so they don’t think they need the medication anymore.” [P16]  “So, it’s the classic thing of, like, someone stops their medication. Therefore, they may relapse, so then there’s a likelihood of, or a higher likelihood of, someone getting referred again.” [P23]  “They think sometimes when they come out of hospital, maybe it’s lack of education that they think, “Oh, I don’t need to take it anymore. I’m not hospital, I’m fine,” when obviously we know that there needs to be continuation with a lot of the medications.” [P2] | “Sometimes some of them (people from black backgrounds) maybe migrate to this place without any family. They do not have any family around; they are not understanding how things work.” [P9]  “Well, his friends and family lived elsewhere, so in a different city, so that was part of the difficulty. So, he didn’t have much in the way of social structure around him.” [P16]  “So, definitely, sometimes people who have less support around them can be a bit more tricky to be in touch with. You might find out, only, a lot later things have progressed a lot more.” [P15]  “Lack of social support, lack of protective factors, so someone becoming- often if someone does have, say, a supportive network, family or friends, it can be intercepted. The relapse will- the worsening of symptoms will be noticed more easily and quickly.” [P14] | "It could also be to do with them also having more factors working against them in terms of deprivation, you know, socioeconomic situation which means that they become unwell more as a result of those discriminatory factors." [P14]  "If you belong to a very high socioeconomic group…then they may be less represented as a section patient in hospital." [P22]  “And this person actually would not come to a clinic because it was a different area and he was worried about the local gangs.” [P13]  “We’re not always very good at reaching out into certain communities, to support understanding, and what kind of services are out there, and how they can access help.. until things are much worse”. [P20] | “Probably the cultural understanding, but their families and relatives can impact that. A lot of families because their culture want to keep it in their community, don’t like asking for support. Some of it could be poverty-deprived and stereotypes in the community.” [P2]  “Most people from – I’m generalising now – ethnic back grounds, black background specifically, sometimes, do believe in this juju, voodoo, all those aspects.” [P9]  “They might not understand that this is a medical condition, rather than a spiritual thing. So, they might be seeking maybe more spiritual help, rather than a combination of both.” [P9] |

| **Theme** | **Service/Clinician Level Factors Contributing to People Being Compulsorily Detained** | | |
| --- | --- | --- | --- |
| **Sub Themes:** | **Lack Of Communication and Continuity in Care** | **Previous Failure to Build Trust** | **Clinician Biases and Assumptions** |
| **Additional Quotes** | “I don’t think necessarily that everyone communicates well with one another. And when people have been discharged from hospital, as you know, there are quite a few different services and people involved in a patient’s care, and I don’t necessarily think that that flow is done properly.” [P2]  “You lose information every time you hand over to a new care coordinator, even though there is written information. You lose that therapeutic relationship.” [P4]  “It’s not very transparent to me what individual people are going to receive. There is not a lot of communication between inpatient and community settings, and that’s part of the problem . I can make recommendations, but I have no control whatsoever. I have no way of following up necessarily, but it’s not my role to follow up, unless I’m asked to. Oftentimes, things are put in place that are just not delivered due to lack of resources.” [P7] | “Somebody from a Black British background has experienced a lot of discrimination in their life, and that can be projected as more likely to distrust authority, and lack of cooperating because of experiences that they’ve had.” [P22]  “Especially in BAME populations it is already too late because there has been an initial non-engagement because there is a bit of a distrust of mainstream services.” [P4]  “They present with more externalised behaviour, and I think it comes from not having trust. I think that if you don’t trust authority or police. I mean when we are scared, we call the police. If you don’t feel the whole system is there to support you, if you are scared or frightened you will act out. So it’s the ability to negotiate with the wider kind of social structure. So you’re immediately marginalised.” [P13] | “I think there’s an unconscious bias still, and a narrative that plays out through healthcare, around the risks of someone who’s black and might be psychotic, I think, which leads to people thinking they may need to be sectioned.” [P20]  “Someone with psychosis is more likely to be admitted under the Mental Health Act, than someone with depression.” [P18]  “If there’s already a history, like back to your original question, there was already history of sectioning, then it feels easier. You would determine: “Well, there’s already a nature, so we can consider this person is – they look – quite poorly. They seem quite aggressive, and they don’t want to engage.” [P23]  "If there’s already a history, like back to your original question, there was already history of sectioning, then it feels easier. You would determine: “Well, there’s already a nature, so we can consider this person is – they look – quite poorly. They seem quite aggressive, and they don’t want to engage." [P23] |

| **Theme** | **Service/Clinician Level Factors Contributing to People Being Compulsorily Detained** | | |
| --- | --- | --- | --- |
| **Sub Themes:** | **Lack Of Resources and Disruptions** | **Lack Of Variety of Treatments/Care Offered Leading To Compulsory Detention** | **Systemic And Institutional Barriers to Engagement** |
| **Additional Quotes** | “As a care coordinator it’s really difficult at the moment just because of the caseloads. So in better times it would be the case that a care plan is done and reviewed regularly, but we are in quite a critical situation at the moment and were not able to do that as much.” [P14]  “Especially with COVID and lots of cutbacks, resourcing has changed (…) I think people are discharged very quickly now.” [P19]  “Huge holes in funding… services have such limited resources, such limited capacity.” [P8]”  “Issues around resources, the lack of community options. There is something about the discharge processes that doesn’t optimise people’s abilities when they’re back in the community.” [P2]  “There are certain people that get sectioned again because the amounts of resources in the community are not there enough to provide a level of intensive support in the community.” [P22] | “I would have wanted us to have much more psychology provision to make space or us to understand psychologically what is going on with this person […] It’s my view that we don’t work in settings that allow us to sit in a room with a person for 45 minutes or an hour to unpick and explore with them what it is that might be going on, there simply isn’t space for that.” [P12]  “[patient getting sectioned again] was partly as a result of us not fulfilling our side of the bargain. Our side of the bargain was that we would ensure that she had psychological therapy as soon as possible.” [P1]  “I think another factor, going back to the medicalised model, is that although drug treatments can be really important and effective for people, especially in a crisis, so really helpful and beneficial short term therapeutic effects, in the longer term those medications create all kinds of problems for people.” [P4] | “The other thing to think about is are there aspects of institutional racism going on here. So, we know that black men are more likely to also be in secure settings and prisoned proportionately as well. So, is there an element of, you know, of that going on in terms of, like, a more broader institutional issue around racism.” [P4]  “I think there’s probably an element of institutionalised racism within mental health systems as well as many other systems in this country.” [P17]  “I think in some cases maybe BME, certain groups of BME people might not want involvement with services because they feel that services are discriminatory or systemic.” [P14] |

| **Theme** | **Ways of Reducing Compulsory Detention** | | |
| --- | --- | --- | --- |
| **Sub Themes:** | **Improving Quality of Care, Continuity of Care, and Communication** | **Increasing Access to Patient & Family Level Intervention** | **Investing in Services** |
| **Additional Quotes** | “It’s also really important that the people who run and deliver services also reflect the ethnic background and the people of the communities that they’re serving as well. So, I would say that was an important issue that we have diversity.” [P4]  “We need to get better at adapting very Eurocentric ways of working, and I make this really clear locally as well in the work that we do. Becoming trauma informed in our practice is a step in the right direction to address that.” [P12]  “When you have good communication, and you make a good plan, and everybody is signed up to it, your chances of success are much greater.” [P16]  “Having capacity to engage with people in a meaningful way. I think just the act of people feeling supported, seen, heard, listened to can reduce the risk of people becoming unwell and reduce the risk of, therefore, people being detained.” [P19]  “So we need to move away from diagnoses, psychologists have got it right, we need a formulation based approach.” [P1] | “We need access to all the other things that can help keep people well and away from psychiatric hospitals. Things like access to effective substance misuse services, which is a big problem for a lot of my patient population.” [P17]  “Working with family is a really important part of what we do, and it happens very little. It can be quite hard to make contact with family. It can also be hard to engage family in what we used to call psychoeducation work, but to get families to come along to sessions.” [P16]  “Getting people linked into community resources is also quite a helpful way of keeping people well.” [P19]  "I think what would really help is the expansion of more preventative services." [P14] | “…if they had more resources, and there were- I guess, if there was more care available to him, it might have helped prevent him getting to the stage where there was no other option.” [P18]  “Wards should be given more time to facilitate someone’s discharge in a very planned way, that accesses them into community resources in a more comprehensive way might be a way to reduce the likelihood [of being sectioned].” [P5] |

| **Theme** | **Ways of Reducing Compulsory Detention** | |
| --- | --- | --- |
| **Sub Themes:** | **Offering Variety of Treatments/Care and Alternatives to Compulsory Detention** | **Improved Discharge Planning** |
| **Additional Quotes** | “Maybe we need more of the expansion of crisis house, there’s always a lack of bedding at crisis houses in the area. They do a really good job at again preventing admission. So it’s more like focusing on preventative respite services.” [P14]  “If we were sometimes less resistant to the idea that- sometimes you see things where you can see the pattern of detention and someone having- the pattern of deterioration, sorry, and a patient having some insight, at the point where perhaps an informal admission or a crisis house might have been a viable option.” [P18]  “If there was a community setting that was, perhaps, a bit more intensive than can be offered without admitting, so not a crisis house, then perhaps that would have been an alternative given that the patient was initially agreeable to restarting the medication.” [P17]  “It’s about having more variety in crisis services.” [P4] | “Having really thought-out, planned discharges which are not dictated by bed pressures, not sudden… I feel like it’s very quick now.” [P20]  “I think, in terms of care planning and discharge planning, there’s something that used to happen, going to an inpatient ward, seeing your patient in a ward round and saying, “Let’s just catch up outside of this ward round for 10 minutes and make a bit of a discharge plan or look at this bit of your care plan?” ….Obviously, there are huge benefits to that.” [P17] |
