## Appendix A for "Exploring Pathways to Compulsory Detention and Ways to Prevent Repeat Compulsory Detentions; Clinician Perspectives"

**Study Title: Exploring pathways to detention in psychiatric hospital and ways to prevent repeat detentions: a qualitative study**

**Staff working in mental health services or voluntary sector Interview Schedule**

**Introduction (following taking consent)**

We are planning to adapt an intervention that showed promise in a previous study, to use in UK. We would like to know about your experience, as someone who works with people under section, about what you think leads to being involuntarily admitted, and ask you about the intervention so that we can make the adapted intervention relevant and acceptable/agreeable to those who have more than one compulsory admission to the ward.

1. **Can you tell me how you work with people who get involuntarily admitted under the Mental Health Act?**

Prompt: if not already explained: do you have a role in the process of people being involuntarily admitted under the Mental Health Act?

2. **Can you tell me (without giving any personal information away) about the most recent person you’ve worked with who got “sectioned” under the Mental Health Act or a recent person who has been sectioned more than once? How do you think this came about?**

Probe for stresses, mental health deterioration, factors associated with services, medication and other treatment.

3. **Do you think anything could have been done to prevent this person being sectioned?**

Probe for before any deterioration started, during their deterioration, at the time of the assessment, any types of service or intervention that they think would help.

4. **In general, why do you think people who have been “sectioned” are at high risk of this happening again?**

Probe for factors to do with illness, factors to do with social situation/demographic characteristics, attitudes to their mental health problems and to treatment (including adherence), relationship with professionals, unmet needs for any kind of care.

5. **People from a Black or Black British background are four times more likely than White British people to be detained under the Mental Health Act in the UK. Why do you think this might be the case? What factors might be at play?**

Prompt for any cultural factors/beliefs affecting treatment, lack of trust with services, access/engagement with services, staff biases/perceptions (unconscious or otherwise)

6. **Is there anything you think could be done in services to reduce the risk of people being sectioned again when this has happened once?**

Prompt for: what the ward could do to reduce risk, what community teams could do to reduce risk, risk assessments, care plans and how they are monitored/used in practice by services, involving family/friends, signposting to community organisations, prompt for own examples

**Then, researcher will give a brief explanation of the intervention:**

The next part of the interview is about the new type of support we are developing. The aim is that this new support will make it less likely that someone who has been sectioned gets sectioned again once they are discharged from hospital.

The new support will involve four one hour sessions delivered by a psychologist starting in the hospital ward.

The psychologist will work with the person to understand what led up to them being sectioned, and identify any signs that could indicate another crisis is developing, and how to respond. The psychologist will help the person to make a plan to better manage their day-to-day mental health when they leave hospital, including thinking about what might help them to stay well and who to contact if their mental health gets worse. This plan will either be in electronic or paper form, and the person can keep and refer to it when they leave hospital.

We would like to take a positive, person-centred approach when delivering this support. The psychologist will focus on who the person is and what their strengths, values and resources are, and help them to set meaningful goals they’d like to pursue when they leave hospital.

After these four sessions, the person will continue to receive support over the course of the next 12 months. The psychologist will schedule a monthly 45 minute call to check in and discuss how the person is doing, and whether they have been using the plan that they created. The plan might need to be adjusted over the course of the year if anything changes once the person has left hospital.

**[Share screen with intervention diagram at this point]**

7. **What do you think about the intervention described?**

Prompt for: positive and negative aspects, What do you most like about it?

For Slide 2 – how would this work in a 4 x one hour sessions? Are there elements you think are more important than others, bearing in mind that the intervention aims to reduce readmission to hospital?

What ideas or suggestions, from your clinical practice for example, do you have about the best way to make an intervention like this appropriate to people from diverse backgrounds.

8. **Is there anything you would change about this intervention?**

Prompt for: Length? Time? Intensity? Mode? Specific elements?

9. **Do you think the psychologist providing this help should keep in touch with the participant’s care coordinator and mental health team? Should this be part of the participant’s community care plans?**

E.g. attending joint meetings, sharing advanced directives with broader team and involving them in any care/crisis plan

10. **We want to make sure that the introductory materials and recruitment strategies for the pilot trial are relevant and engaging, and to ensure ward staff and patients clearly understand the purpose of the intervention.**

**What useful materials could we give participants and staff to help them understand how this intervention will work and why we are doing it?**

Prompt for: how to make intro materials more accessible/engaging (e.g. infographics), explaining key terms to people e.g. “randomization”, Participant Information Sheets can be very long – how to condense key information and display it?

How can we encourage staff to help us recruit people on the wards? Best ways to engage with busy ward staff?

11. **How do you think we could make it easier to get people interested in trying our approach out?**

Prompt for: ways to aid understanding, ways to increase motivation, best ways to engage and build trust between us as researchers and people on the ward, how to distinguish between our intervention and other therapies on the ward?

**End of questions**
